# Urine Test Predicts Kidney Injury and Death in COVID-19

**DOI:** 10.1101/2021.06.10.21258638

**Authors:** Katherine Xu, Ning Shang, Abraham Levitman, Alexa Corker, Satoru Kudose, Andrew Yaeh, Uddhav Neupane, Jacob Stevens, Sumit Mohan, Rosemary Sampogna, Vivette D’Agati, Krzysztof Kiryluk, Jonathan Barasch

**Affiliations:** Department of Medicine, Columbia University, New York; Department of Pathology, Columbia University, New York; Department of Epidemiology, Columbia University, New York

## Abstract

**Background:** Kidney injury is common in COVID-19 infection, but serum creatinine (SCr) is not a sensitive or specific marker of kidney injury. We hypothesized that molecular markers of tubular injury could diagnose COVID-19 associated kidney damage and predict its clinical course.

**Methods:** This is a prospective cohort study of 444 consecutive COVID-19 patients (43.9% females, 20.5% African American, 54.1% Latinx) in Columbia University’s Emergency Department at the peak of the New York pandemic (March-April 2020). Urine and blood were collected simultaneously at admission (median time of day 0, IQR 0-2 days) and within 1 day of a positive SARS-CoV-2 test in 70% of patients. Biomarker assays were blinded to clinical data.

**Results:** Urinary NGAL (uNGAL) was strongly associated with AKI diagnosis (267±301 vs. 96±139 ng/mL, P=1.6×10^−10^). uNGAL >150ng/mL had 80% specificity and 75% sensitivity to diagnose AKIN stage 2 or higher. uNGAL quantitatively predicted the duration of AKI and outcomes, including death, dialysis, shock, and longer hospital stay. The risk of death increased 73% per standard deviation of uNGAL [OR (95%CI): 1.73 (1.29-2.33), P=2.8×10^−4^] and was independent of baseline SCr, co-morbidities, and proteinuria [adjusted OR (95%CI): 1.51 (1.10-2.11), P=1.2×10^−2^]. Proteinuria and uKIM-1 also indicated tubular injury but were not diagnostic of AKI. Typically, distal nephron segments transcribe NGAL, but in COVID-19 biopsies with widespread acute tubular injury (ATI), NGAL expression overlapped KIM-1 in proximal tubules.

**Conclusion:** uNGAL predicted diagnosis, duration, and severity of AKI and ATI, as well as hospital stay, dialysis, shock, and death in patients with acute COVID-19.

## Introduction

Acute loss of kidney function, measured by a rise in serum creatinine (SCr), is common in the setting of acute SARS-CoV-2 infection^1–4^. Elevated SCr is present in one-third of patients hospitalized with COVID-19^5–11^. Yet, SCr fails to represent the full burden of COVID-19 kidney injury, because it cannot diagnose early stages or subtotal kidney damage^12,13^. Moreover, a rise in SCr does not reveal the anatomical or physiological basis of kidney dysfunction. For example, an isolated SCr level cannot distinguish volume depletion from tubular injury, the two common entities that contribute to renal manifestations of COVID-19 and which must be distinguished for appropriate triage and fluid management at hospital admission^10^. Therefore, the well-established association between COVID-19 and loss of kidney function begs the question as to the best noninvasive method to detect COVID-19 associated tubular injury, to identify the mechanisms of functional failure, and to prognosticate the outcome of the illness.

Prior research in human and mouse models has demonstrated that two molecular markers of tubular injury, urinary NGAL (uNGAL) and urinary KIM-1 (uKIM-1) derive from different segments of the kidney and allow for sensitive detection and real time distinction between volume depletion and acute tubular injury (ATI)^12,14–16^. Consequently, these biomarkers may elucidate the pathogenesis of COVID-19 associated kidney injury, but have not been tested in this setting. Here, we assessed their performance in the diagnosis and stratification of kidney injury in acute COVID-19 patients presenting to Columbia University’s Emergency Department at the peak of the New York City pandemic (March-April, 2020). We tested uNGAL and uKIM-1 for association with the diagnosis, duration, and severity of Acute Kidney Injury (AKI) defined by SCr-based AKIN criteria (primary outcomes), as well as with in-hospital death, dialysis, shock, respiratory failure, and length of hospital stay (secondary outcomes).

## Materials and Methods

### Columbia University COVID-19 Biobank and Study Cohorts

The Columbia University COVID-19 Biobank was established in response to the surge of SARS-CoV-2 in New York City in March 2020. The Biobank recruited consecutive COVID-19 cases, regardless of age, sex, or race/ethnicity, diagnosed and treated at Columbia University Irving Medical Center (CUIMC). All included cases were diagnosed by positive SARS-CoV-2 PCR test based on nasopharyngeal sampling. The biobank collected residual blood and urine samples after clinical testing from every COVID-19 patient diagnosed at CUIMC, providing us with the largest cohort of COVID-19 patients for urinary biomarker analysis to date^17,18^.

The cohort was recruited from 3/24/2020 to 4/27/2020 with exclusion only for end stage renal disease on dialysis (Supplemental Figure S1) and linked to the Electronic Health Records for patient characteristics and outcomes (Supplemental Table S1). Shock was defined by the need for vasopressors, and respiratory failure was defined by the need for either invasive or non-invasive positive pressure ventilation.

Kidney biopsies were accessioned by the CUIMC Renal Pathology Laboratory from 3/13/2020 to 6/1/2020, including 13 kidney biopsies from COVID-19 cases^19^ and 4 non-COVID-19 specimens.

### AKI definitions, AKIN severity stages, and AKI sub-stratification

Baseline SCr was determined, as described by Stevens et al.^20^, as follows in order of preference,

- Median SCr from 365 to 31 days before urine collection. If not available, then:
- Minimum SCr from 30 days before urine collection to the day of collection. If not available, then:
- Minimum SCr from urine collection to 7 days in hospital.

The patient was classified as “unknown” status if none of the above criteria were met.

The loss of kidney excretory function was determined by SCr kinetics, according to Acute Kidney Injury Network (AKIN) definitions^21^, and interpreted as an absolute increase in SCr of ≥0.3mg/dL or a ≥ 50% increase from baseline. AKIN stages were classified as follows:

- AKIN Stage 1: ≥0.3mg/dL increase in sCr within a 48-hour window OR 1.5 to 2-fold increase in sCr compared to baseline.
- AKIN Stage 2: >2 to 3-fold increase in SCr compared to baseline.
- AKIN Stage 3: ≥0.5mg/dL increase in SCr within a 48-hour window when SCr ≥4.0mg/dL OR >3-fold increase in SCr compared to baseline.

The first SCr in days 0-2 after urine collection was used to diagnose AKIN stage using the above criteria. In select cases, the day 1 AKIN score was imputed when the preceding and subsequent AKIN scores were identical. Further categorization of elevated SCr was based on the duration of SCr elevation above baseline:

- No AKI (AKIN=0) – not meeting AKIN criteria within 2 days of presentation (must have SCr values for both days).
- Transient AKI (tAKI) – met AKIN criteria on day 0 or 1 of presentation but normalized below AKIN detection thresholds within 2 days after first detection (total AKI duration <72 hours).
- Sustained AKI (sAKI) – met AKIN criteria within 2 days of presentation but normalized below the AKIN detection thresholds only after 2 days from the first detection (total AKI duration >72 hours).
- Unknown – missing baseline SCr, or insufficient SCr measurements to determine SCr kinetics, or missing measurements on day 0 or 1 that could not be imputed due to discrepant AKIN scores.

Diagnosis and staging of chronic kidney disease (CKD) utilized KDIGO criteria^22^.

### Urinary biomarker assays

uNGAL and uKIM-1 were measured by ELISA (NGAL: BioPorto, KIT036, KIM-1: Enzo, ADI-900-226-0001) according to manufacturer’s protocols. Proteinuria levels were detected using Chemstrip 10 SG urine test strips (Roche Diagnostics). Measurements of uNGAL, uKIM-1, and proteinuria were blinded from clinical data.

### Urinary cell pellet Western blots

Urinary cell pellets were analyzed for LRP2 and UMOD by immunoblot with SDS-PAGE (Bio-rad Laboratories), rabbit anti-LRP2 (1:1000, Abcam, ab76969), sheep anti-UMOD (1:2000, Meridian Life Science, K90071C), and polyclonal secondary antibodies conjugated to HRP (1:10,000, Jackson Immuno-Research).

### uNGAL dipstick measurements

Urine was centrifuged (12,000rpm; 10min) and 10uL applied to NGAL gRAD dipsticks (BioPorto). Color development over 15min was compared to the manufacturer’s test line by two independent, blinded readers and associated with semi-quantitative bins for risk of kidney damage.

### Pathological Analysis

Extent of acute tubular injury (ATI) was assigned based on the following semi-quantitative scale: none (<5% of tubules involved), mild (<25%), moderate (25-50%), severe (>50%). Histologic features of ATI included loss of brush border, epithelial simplification, intracytoplasmic vacuolization, overt necrosis, apoptosis and cell shedding.

### Renal Ischemia Reperfusion Injury Model

Male and female wild-type C57Bl/6 mice, aged 8-10 weeks (Jackson Labs) were anesthetized with isoflurane and placed on a warming table to maintain a rectal temperature of 37°C. The left renal pedicle was clamped for 10, 20, 30, or 40 minutes using microvascular clamps (Fine Science Tools). After the clamps were removed, reperfusion of the kidneys was visually confirmed. The kidneys were harvested at 24hrs.

### RNAscope® In Situ Hybridization (ISH)

In Situ Hybridization (ISH) on formalin-fixed paraffin-embedded (FFPE) human kidney tissues was performed using the chromogenic RNAscope® 2.5 HD Duplex Reagent Kit (Advanced Cell Diagnostics, 322430) according to the manufacturer’s protocols. The following probes were used in dual channels: Hs-LCN2 (559441), Hs-LCN2-C2 (559441-C2 at 1:600), Hs-HAVCR1-O1 (538081 and 538081-C2 at 1:100), Hs-AQP2 (434861), and Hs-LRP2 (532391). Mouse sections were probed with Mm-Lcn2-C2 (313971-C2) and Mm-Havcr1 (472551). Tissue sections were counterstained with hematoxylin. Bright field images were captured using the Olympus IX73 Inverted Microscope under low (100x) and high (600x) magnifications.

### Statistics

Continuous variables were compared using a two-sample t-test and summarized as mean ± SD. Non-normally distributed continuous variables (i.e., uNGAL, uKIM-1) were natural log-transformed and standard-normalized before statistical testing. Categorical variables were compared using Chi-squared or Fisher’s Exact test. For testing binary outcomes, we used logistic regression. Ordinal outcomes, such as AKIN stage, were tested using ordinal logistic regression. Ordinal predictors, such as urine dipstick category or proteinuria grade, were tested under the assumption of linear effects using a slope test within the framework of a generalized linear model tailored to the outcome of interest (e.g. logistic or ordinal logistic for binary or ordinal outcomes, respectively). We used Cox proportional hazards model for the time-to-event analyses of mortality. We used competing risks regression model for the analysis of the length of hospital stay, with death as a competing risk^23,24^. The proportional hazards assumption was verified by testing scaled Schoenfeld residuals for each predictor against observation time. Associations of urinary biomarker levels with clinical outcomes were adjusted for the following covariates: age, sex, race, and ethnicity (minimally-adjusted model), baseline serum creatinine and pre-existing obesity, diabetes, hypertension, transplant (any organ), cancer (hematologic cancers and any solid tumors), cardiovascular disease (coronary artery disease, heart failure, cerebral infarction), pulmonary disease (asthma, chronic obstructive pulmonary disease, interstitial pulmonary disease, primary pulmonary hypertension, idiopathic pulmonary fibrosis) (fully-adjusted model-1), as well as proteinuria (fully-adjusted model-2). In the analysis of primary outcomes, we considered two-sided P<0.05 as statistically significant. In the analysis of secondary outcomes, we considered P<0.01 as significant, corresponding to the Bonferroni-correction for 5 major independent outcomes tested. All statistical analyses were performed using R (CRAN version 4.0.4), including R add-on packages MASS (version 7.3-53.1), odds ratio (version 2.0.1), survival (version 3.2-7), cmprsk (version 2.2-10), and pROC (version 1.17. 0.1).

### Oversight of Human and Animal Studies

The Columbia University Biobank COVID-19 studies were reviewed and approved by the Columbia University Medical Center Institutional Review Board (IRB). Samples and tissues were collected and stored in the Columbia University COVID-19 Biobank in accordance with the Institutional Review Board of Columbia University protocol (IRB AAAS7370) while the study of kidney injury biomarkers was approved by the IRB AAAS7948. A subset of patients was included under a public health crisis IRB waiver of consent specifically for COVID-19 studies if patients were deceased, not able to consent, or if the study team was unable to contact them as per the Columbia Institutional Review Board protocols. Mice were utilized according to the Institutional Animal Care and Use Committee (AC-AAAY7464) and adhere to NIH Guide for the Care and Use of Laboratory Animals. Our study was reported according to STROBE guidelines for cohort studies^25^.

## Results

We analyzed urine samples from 444 COVID-19 patients collected prospectively by the Columbia University COVID-19 Biobank at a median time of day 0 (hospital admission, IQR 0-2 days), within 1 day of a positive SARS-CoV-2 test in 70% of patients (Supplemental Figure S1). The cohort was diverse in age, sex, race, ethnicity, and pre-existing comorbidities (Supplemental Table S1).

Admission uNGAL levels were higher among patients reaching the diagnosis of AKI (uNGAL: 267±301 vs. 96±139 ng/mL, P=1.6×10^−10^) or sustained AKI (lasting >72 hours; sAKI; uNGAL: 332±324 vs. 96±139 ng/mL, P=3.3×10^−11^) compared to those without AKI (Figure 1a). uNGAL levels were also associated with increasing AKIN stage (slope P=4.0×10^−21^) (Figure 1b). Accordingly, the area under the receiver operating characteristics (ROC) curve for uNGAL increased for higher AKIN stages (Figure 1c). For example, uNGAL had 80% specificity and 75% sensitivity to diagnose AKIN stage 2 or 3 at a cutoff level of 150ng/mL (Table 1). Notably, uNGAL levels were marginally elevated in patients who experienced transient AKI (tAKI) (187±257 vs. 96±139 ng/mL, P=0.021) or the AKIN-1 stage (162±219 vs. 96±139 ng/mL, P=6.8×10^−3^) compared to those with no AKI.

**Figure 1.**
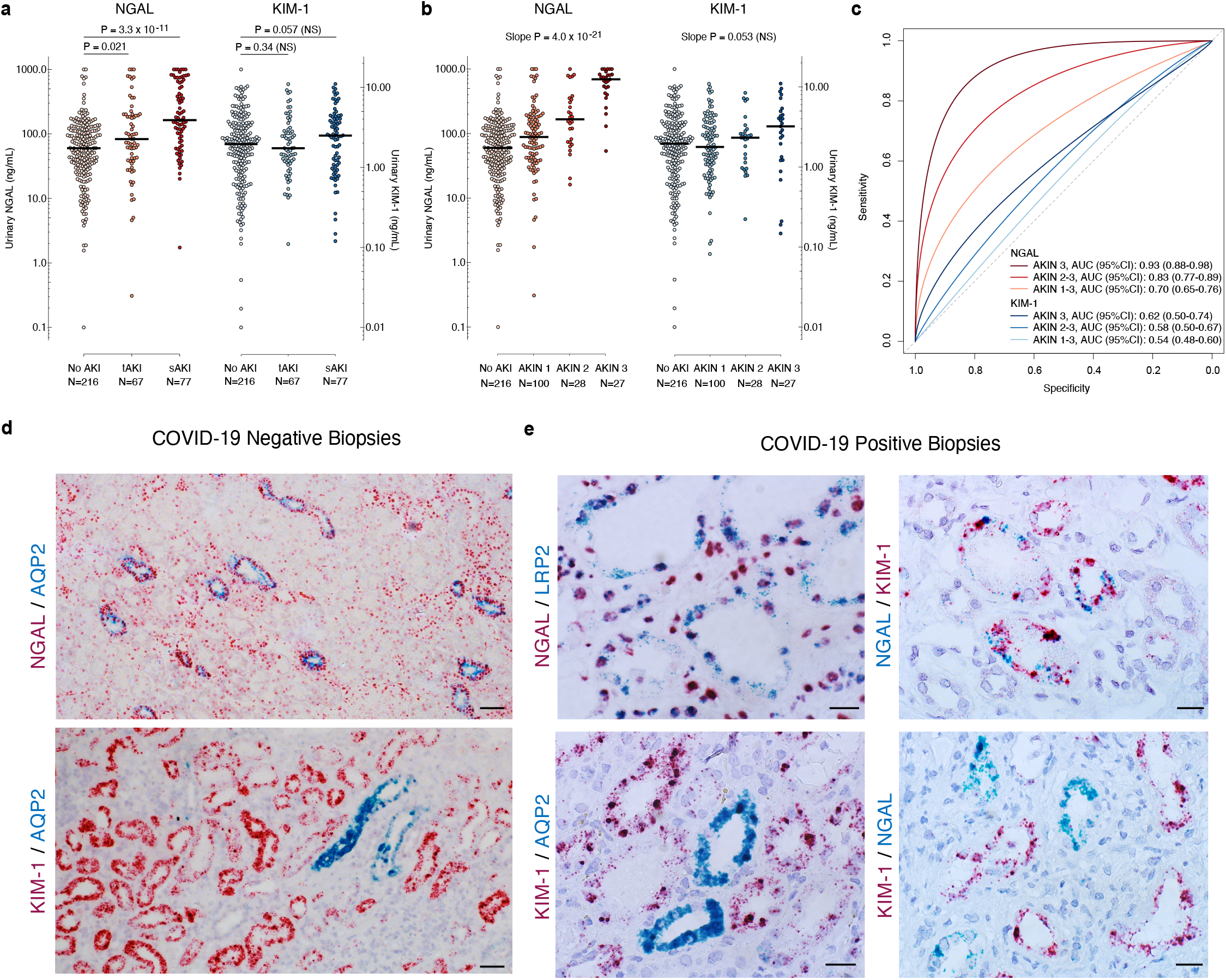
uNGAL is associated with the duration and severity of acute tubular injury in COVID-19 patients: **(a)** uNGAL, but not uKIM-1, was associated with sustained AKI (sAKI, meeting AKIN criteria for ≥72hrs) in COVID-19 patients. No AKI and transient AKI (tAKI, <72hrs) levels are shown for comparison. **(b)** uNGAL, but not uKIM-1, was associated with the severity of AKI (AKIN stage); bars represent medians. Notably, mean levels of uKIM-1 were equally elevated in all four groups, including in COVID-19 patients with AKIN stage 0 (no elevation of SCr). **(c)** ROC curves for uNGAL (shades of red) and uKIM-1 (shades of blue), by ascending AKI severity (AKIN 1-3 vs. 0, AKIN 2-3 vs. 0-1, AKIN 3 vs. 0-2). **(d)** In non-COVID-19 AKI biopsies, NGAL (*LCN2*) mRNA is expressed in the distal nephron including AQP2+ collecting ducts (top panel), while KIM-1 (*HAVCR1*) is expressed in proximal tubules and not in AQP2^+^ collecting ducts (bottom panel). Scale bars=50µM. **(e)** COVID-19 positive kidney biopsies with widespread acute tubular injury (80-100% of tubules) demonstrated extensive expression of NGAL in a non-canonical distribution, in LRP2^+^ and KIM-1^+^ proximal tubules (top panels), whereas COVID-19 kidney biopsies with limited ATI (30% of tubules) exhibited limited NGAL-KIM-1 overlap (bottom right panel). KIM-1 was expressed only in AQP2^-^ proximal tubules (bottom left panel). Scale bars=20µM.

**Table 1.** Diagnostic properties of urinary NGAL test at test cutoffs corresponding to specificity of 80%, 85%, and 90%. N=371. PPV, positive predictive value; NPV, negative predictive value; +LR, positive likelihood ratio; -LR, negative likelihood ratio.

| **Outcome** | **Cutoff**<br>**(ng/mL)** | **Cutoff**<br>**(z-score)** | **Specificity** | **Sensitivity** | **PPV** | **NPV** | **+ LR** | **- LR** |
| --- | --- | --- | --- | --- | --- | --- | --- | --- |
| Sustained AKI | 150 | 0.50 | 0.80 | 0.57 | 0.44 | 0.87 | 2.88 | 0.53 |
|  | 175 | 0.60 | 0.85 | 0.49 | 0.48 | 0.86 | 3.31 | 0.60 |
|  | 205 | 0.71 | 0.90 | 0.45 | 0.56 | 0.86 | 4.58 | 0.61 |
| AKIN 1-3 | 130 | 0.40 | 0.80 | 0.51 | 0.65 | 0.69 | 2.55 | 0.61 |
|  | 150 | 0.50 | 0.85 | 0.47 | 0.70 | 0.69 | 3.16 | 0.62 |
|  | 170 | 0.58 | 0.90 | 0.43 | 0.75 | 0.68 | 4.16 | 0.64 |
| AKIN 2-3 | 150 | 0.50 | 0.80 | 0.75 | 0.39 | 0.95 | 3.67 | 0.32 |
|  | 172 | 0.59 | 0.85 | 0.71 | 0.45 | 0.94 | 4.65 | 0.34 |
|  | 207 | 0.72 | 0.90 | 0.64 | 0.52 | 0.93 | 6.26 | 0.40 |
| AKIN 3 | 163 | 0.55 | 0.80 | 0.93 | 0.27 | 0.99 | 4.60 | 0.09 |
|  | 185 | 0.64 | 0.85 | 0.93 | 0.33 | 0.99 | 6.23 | 0.09 |
|  | 234 | 0.80 | 0.90 | 0.89 | 0.41 | 0.99 | 8.71 | 0.12 |

Measurement of uNGAL using a novel rapid point-of-care semi-quantitative dipstick^20^ strongly correlated with ELISA measurements (Spearman’s correlation ρ=0.84, P=1.5×10^−119^), reproducing the association of uNGAL with AKI, sAKI, and AKIN stages at the same level as ELISA uNGAL (Supplemental Figure S2).

The association of uNGAL with the primary outcomes was independent of age, sex, race and ethnicity (minimally-adjusted model), baseline creatinine and other comorbidities (fully-adjusted model-1) and was also independent of proteinuria measured in the same urine sample (fully adjusted model-2; Supplemental Table S2). In addition, uNGAL was quantitatively associated with four out of five secondary outcomes of COVID-19, including in-hospital death, acute dialysis, shock, and length of hospital stay (Figure 2a), independent of demographics, comorbidities, baseline renal function, and proteinuria (Supplemental Table S2).

**Figure 2.**
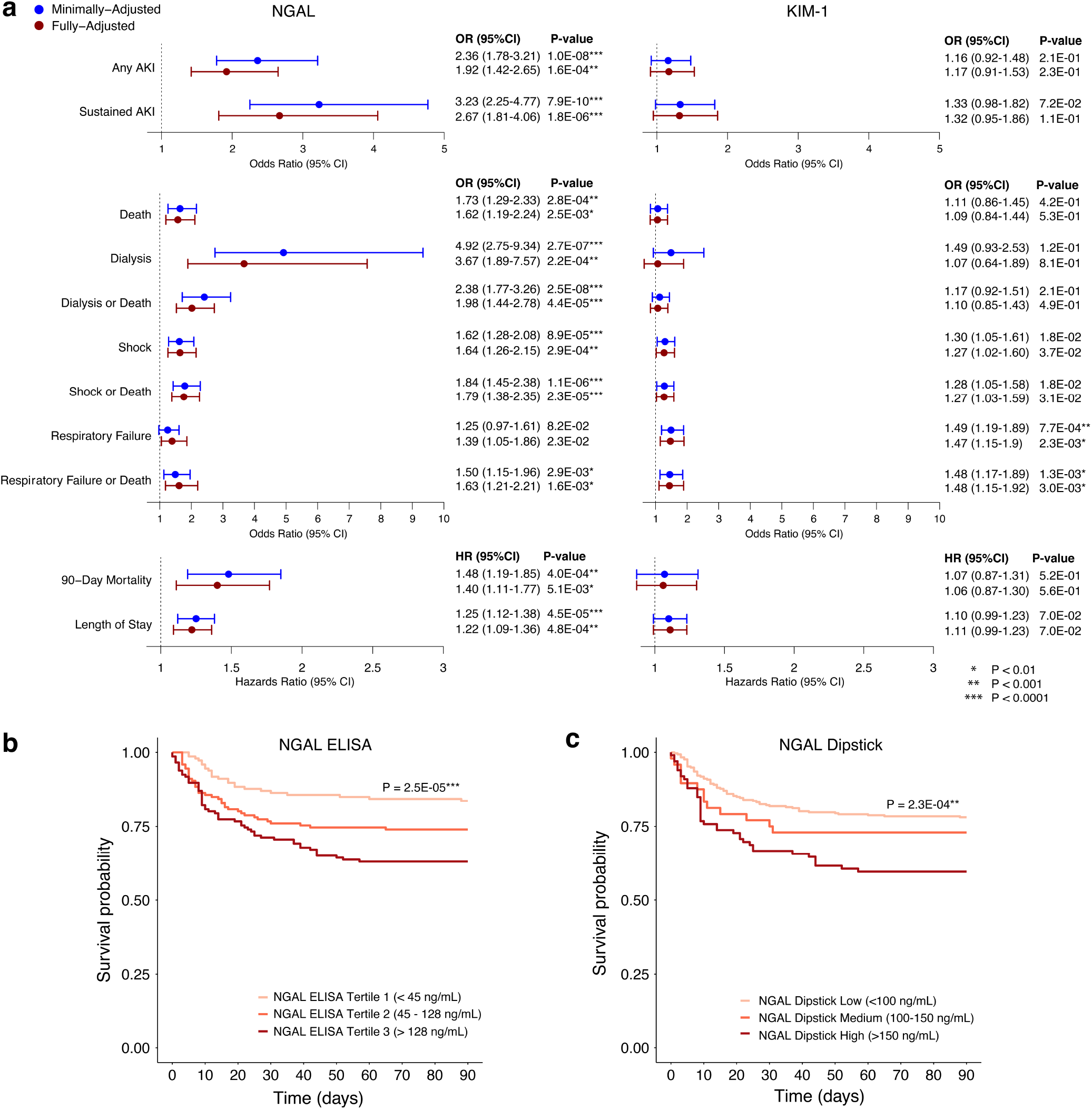
Higher urinary NGAL levels are associated with critical illness and death in patients with COVID-19: **(a)** Urinary NGAL levels were associated with AKI and sustained AKI (>72hrs) after adjustment for age, sex, race, and ethnicity (minimally-adjusted model, blue) and baseline SCr and pre-existing comorbidities (fully-adjusted model, red; N=371). Urinary NGAL levels were also associated with secondary outcomes of death, dialysis, shock, and respiratory failure in both minimally- and fully-adjusted models, N=440. In contrast, uKIM-1 was not associated with AKI or any secondary outcomes except for respiratory failure. Odds ratios (OR) and hazard ratios (HR) are expressed per one unit of standard deviation of biomarker distribution; 95% CI: 95% confidence intervals. **(b)** Kaplan-Meier survival analysis demonstrates survival differences by tertile of urinary NGAL levels measured by ELISA or **(c)** by three levels of urinary NGAL dipstick test (unadjusted P-values provided for both b and c; N=440).

Importantly, uNGAL levels represented an independent predictor of in-hospital death. The risk of death increased 73% per standard deviation of uNGAL [OR (95%CI): 1.73 (1.29-2.33), P=2.8×10^−4^] and this effect was independent of baseline SCr, demographics, or pre-existing co-morbidities [OR (95%CI): 1.62 (1.19-2.24), P=2.5×10^−3^], as well as proteinuria [OR (95%CI): 1.51 (1.10-2.11), P=1.2×10^−2^]. In a time-to-event analysis, the dose-dependent association of uNGAL with 90-day mortality was observed for both ELISA and dipstick measurements. Patients with the highest uNGAL ELISA tertile (>128ng/mL) or the highest uNGAL dipstick category (>150ng/mL) had ∼25% mortality within 20 days of admission (Figure 2b, c).

In contrast to uNGAL, uKIM-1 was not significantly associated with primary outcomes of AKI, sAKI or AKIN stage (Figure 1) nor with secondary outcomes including 90-day mortality or other attributes of critical illness (Figure 2a, Supplemental Table S2).

To determine if our findings were specific to COVID-19, we examined a second cohort of comparable size (426 patients) admitted through the same Emergency Department (6/2017-1/2019)^20^ and analyzed using identical methods (Supplemental Table S1). The COVID-19 cohort was older and enriched in Latinx patients, but the burden of chronic kidney disease was similar in both cohorts. Notably, COVID-19 patients were 2.6-times more likely to present with AKI (35.2% vs. 13.6%, P=2.8×10^−13^), 3.9-times more likely to have sustained AKI (17.5% vs. 4.5%, P=2×10^−9^) and 1.8-times more likely to have more severe disease (AKIN 2-3, 12.5% vs. 6.8%, P=6.6×10^−3^) compared to our historical cohort (Supplemental Table S1), and similar to published data^26^. Urinary findings also differed; both KIM-1 and proteinuria were elevated in all COVID-19 cases compared to the historical cohort, even in patients without AKI, or AKIN=0 (uKIM-1: 2.57±2.44 COVID-19^+^ vs. 1.96±2.51 ng/mL COVID-19^-^, P=7.7×10^−3^; proteinuria severity by ordinal comparison: P=1.3×10^−9^; Supplemental Figures S3 and S4). Consistently, urinary shedding of proximal tubule cells specifically marked by LRP2+ at AKIN=0 was more prominent in COVID-19 than in non-COVID-19 patients (n=40; 2.61-fold increase in urinary LRP2+ cells, P=5.5×10^−3^) while UMOD+ cells were present regardless of COVID-19 status (Supplemental Figure S5). Nonetheless, KIM-1 only marginally increased with AKIN stage in the COVID-19 cohort, while uNGAL quantitatively reflected AKIN stage in both cohorts (Supplemental Figure S3).

To further explore injury in different nephron segments, we examined the transcriptomic patterning of the biomarkers in kidney biopsies from 13 COVID-19 patients^19^ and 4 non-COVID-19 controls with acute tubular injury. In both COVID and non-COVID-19 biopsies, KIM-1 was expressed in the proximal tubule while NGAL was prominently expressed in limbs of Henle and collecting ducts. The distributions were confirmed with segment-specific markers, LRP2 (proximal tubule) and AQP2 (collecting duct). However, in COVID-19 biopsies, NGAL transcripts were widespread. At maximum ATI (>50% of tubules), KIM-1 was expressed in 27% (3,322/12,123), whereas NGAL was expressed in 65.7% (6,580/10,111) of tubules, including significant co-expression in the proximal tubule with KIM-1 in 85% and LRP2 in 77% of kidneys (Figure 1d, e). The NGAL-KIM-1 overlap was associated with the extent of ATI (62.5% vs. 20% of tubules; P=0.02). Similar patterning was induced in dose-response with arterial ischemia in mouse kidney ATI (Figure 3). NGAL RNA expression included the entirety of the cortico-medullary junction, the medulla, and the papilla, as well as the proximal tubule where NGAL overlapped KIM-1. These findings are consistent with a recent report of single cell sequencing after severe ischemic injury^27^.

**Figure 3.**
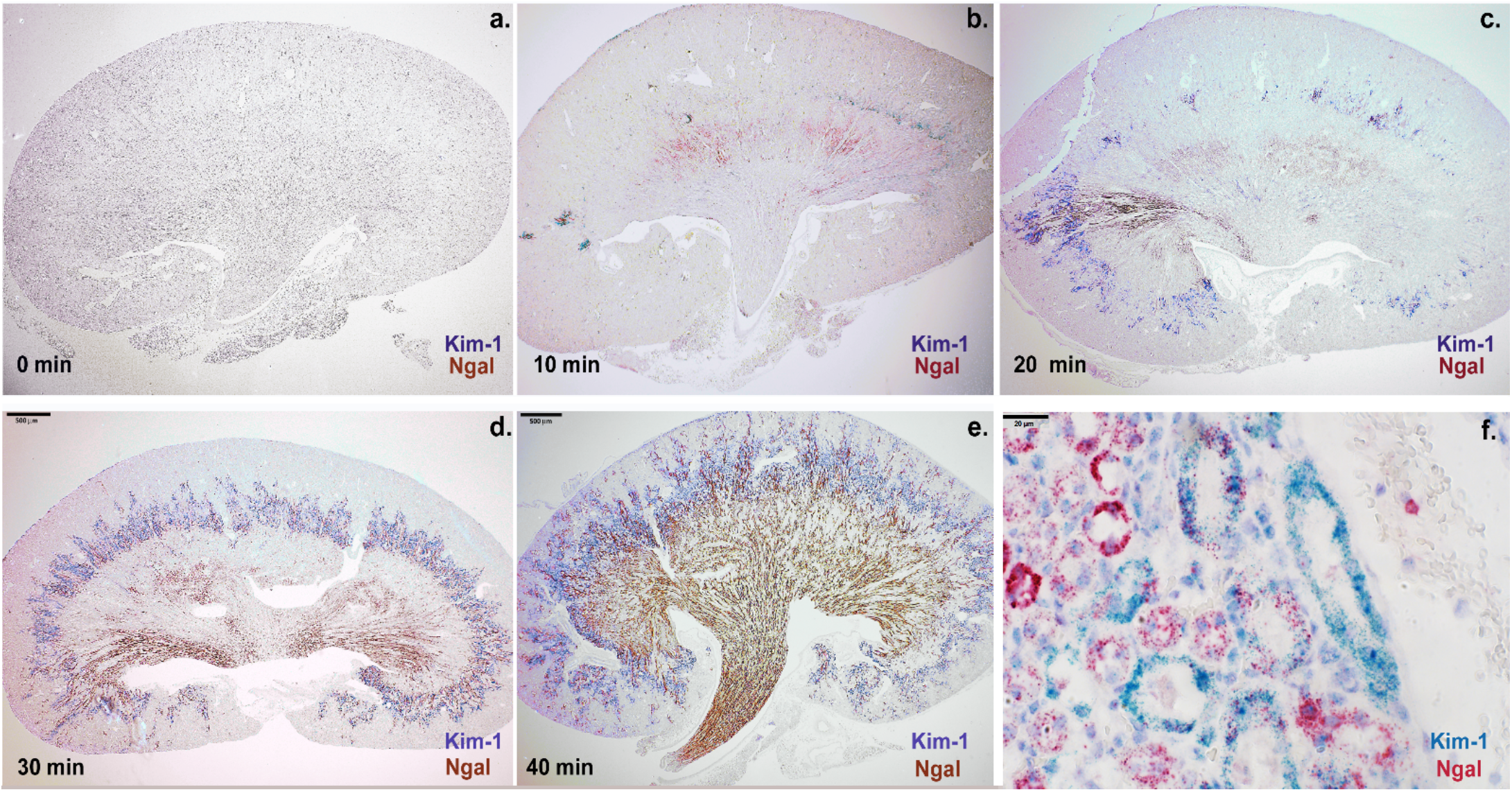
The expression level of Ngal (red-brown) and Kim-1 (blue-purple) depended on the dose of arterial ischemia in mouse: **(a-e)** Ngal expression was found at the cortico-medullary junction after 10min of ischemia, but throughout the medulla and papilla after 30-40min of ischemia. Kim-1 expression was found in the cortex and throughout the cortico-medullary junction. **(e-f)** Prolonged ischemia (40min) broadened the expression domain of Ngal to include the proximal tubule marked by Kim-1. In contrast to Ngal, Kim-1 expression remained localized to the cortex and cortico-medullary junction. Bars a-e: 500μm; Bars f: 20μm.

## Discussion

A simple urine test for NGAL – a protein first discovered in mouse models^13^ – was quantitatively associated with the diagnosis, the severity, and the duration of COVID-19 associated functional kidney failure. Increasing the severity of injury in COVID-19 human kidneys and in mouse models of ischemia broadened NGAL RNA expression. In this light, we demonstrate that the level of uNGAL mirrors the degree of histopathological ATI.

The diagnostic potential of uNGAL to reflect ATI and subsequent functional failure differs from SCr, which increases only as a marker of functional failure^12^. Unlike uNGAL, an acute rise of SCr is delayed after the onset of injury^28,29^ and blunted by mechanisms that enhance its excretion (the renal reserve) or limit its production^30^. In fact, in this study SCr measurements underestimated the impact of COVID-19 since both uKIM-1 and proteinuria were markedly elevated even in the absence of functional kidney failure by SCr criteria (AKIN=0). The shedding of proximal tubule cells specifically marked by LRP2 even at AKIN=0 further demonstrated the insensitivity of SCr to appropriately diagnose proximal tubular injury.

These results provide the possibility of a sensitive diagnostic strategy that bypasses the delays and insensitivity of SCr. Increased proteinuria and uKIM-1 in patients with acute COVID-19 are indicative of proximal tubular injury, while uNGAL, but not uKIM-1, predicts subsequent stages of nephron injury resulting in functional failure and associated clinical outcomes. We suggest that together these biomarkers offer sensitive, timely, and comprehensive evaluation of both tubular injury and functional loss in COVID-19. Moreover, accurate testing is possible at the bedside using a rapid point-of-care dipstick^20,31^, which mitigates the risk of handling infectious body fluids and may be particularly helpful in the setting of high patient volumes witnessed in Emergency Departments during recent COVID-19 surges.

Our study is limited by the use of SCr as the gold standard for AKI diagnosis, since we encountered missing baseline (no records) or follow up (early death or discharge) measurements of SCr in 69 (15.5%) patients. In addition, while we sought rapid diagnosis of COVID-19 associated kidney damage at the time of hospital admission, we were not able to collect urine samples on subsequent days, precluding comparative studies of urinary biomarker kinetics. A recent publication by Nugent et al.^32^ pointed to the need for prolonged patient follow up to determine residual injury of the kidney and to mitigate chronic forms of AKI, and subsequent measurements of uNGAL and uKIM-1 during hospitalization could have provided additional prognostic information. Nonetheless, the level of uNGAL on hospital admission correlated strongly with strict AKIN definitions and identified patients at highest risk of critical illness and death independently of other established risk factors.

In summary, uNGAL is diagnostic of COVID-19-associated AKI, correlates with the severity and duration of AKI, and predicts key clinical outcomes including length of stay, dialysis, shock, and death.

## Data Availability

All data analyses and results are described in this article and supplementary files.

## Author Contributions

JB and KK conceived and designed the study. The Columbia University COVID-19 Biobank collected residual blood and urine samples after clinical testing from every COVID-19 patient diagnosed at CUIMC. KX, JB, UN conducted laboratory measurements including urinary biomarker and protein measurements and in situ hybridizations. NS implemented electronic AKI definitions and analyzed the Electronic Health Records to define clinical outcomes and covariates. AL, AC, AY, KX, JB quantified the distribution of RNA expression. RVS performed mouse experiments. VDA and SK analyzed human kidney biopsy tissue to diagnose AKI and quantify tubular injury. JS and SM contributed to the comparative analysis of COVID-negative dataset. KX and KK performed statistical analyses. KX, JB, and KK wrote the manuscript draft and all authors reviewed and edited the manuscript.

## Acknowledgements

The authors would like to thank all patients for their participation, and the following members of the Columbia University COVID-19 Biobank: Sheila O’Byrne, Renu Nandakumar, Amritha Menon, Yat So, Danielle Pendrick, Eldad Hod, Soumitra Sengupta, Wendy Chung, and Muredach Reilly. The Columbia University COVID-19 Biobank is supported by the Vagelos College of Physicians & Surgeons Office for Research, Precision Medicine Resource (PMR), and Biomedical Informatics Resource (BMIR) of the Columbia University Irving Institute for Clinical and Translational Research (Columbia CTSA).

