## Supplemental Materials for "Urine Test Predicts Kidney Injury and Death in COVID-19"

**Supplemental Figures:**

Figure S1. STARD Diagram: Urine sample collection……………………..…………………………… Page 2
Figure S2. Comparison of urine NGAL measurements by ELISA or Dipstick……………………….. Page 3
Figure S3. Urinary NGAL and KIM-1 in COVID-negative and COVID-positive cohorts ……………... Page 4
Figure S4. Comparison of proteinuria in COVID-19-negative and COVID-19-positive cohorts ..….. Page 5
Figure S5. Shedding of LRP2+ proximal tubule cells into the urine in COVID-19 cases …………... Page 6

**Supplemental Tables:**

Table S1. Baseline characteristics and outcomes for COVID-19 positive and negative cohorts.......Page 7
Table S2. Urinary biomarker associations with primary and secondary outcomes……………........Page 8

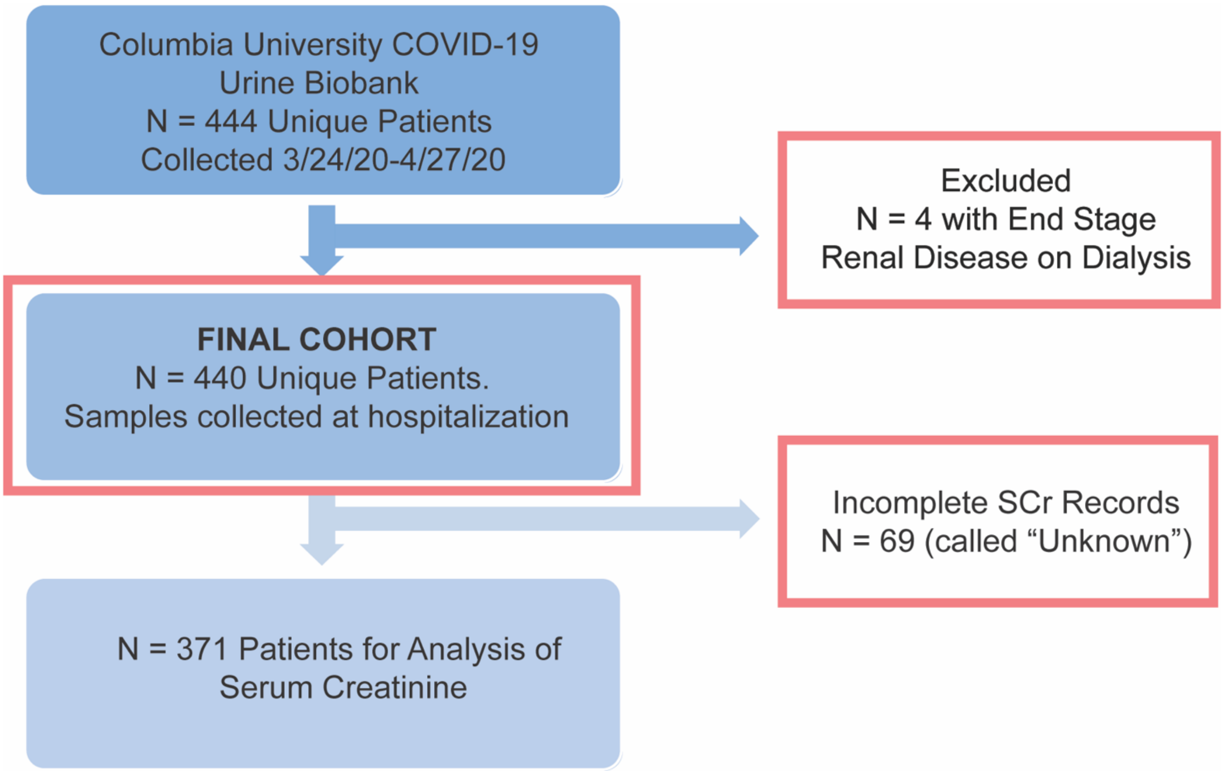


**Figure S1. STARD Diagram: Urine sample collection:** Residual urine samples were collected from 444 consecutive COVID-19 patients by the Columbia University COVID-19 Urine Biobank between 3/24/20 to 4/27/20. Four patients with end stage kidney disease on dialysis were excluded from the final cohort (N=440). Patients with adequate measurements of serum creatinine (SCr) were staged by AKIN criteria (N=371) and subtyped by AKI duration as transient (elevation in SCr < 72 hours) or sustained (elevation in SCr ≥ 72 hours) AKI. Sixty-nine patients missing adequate kidney function data were labeled “Unknown.”

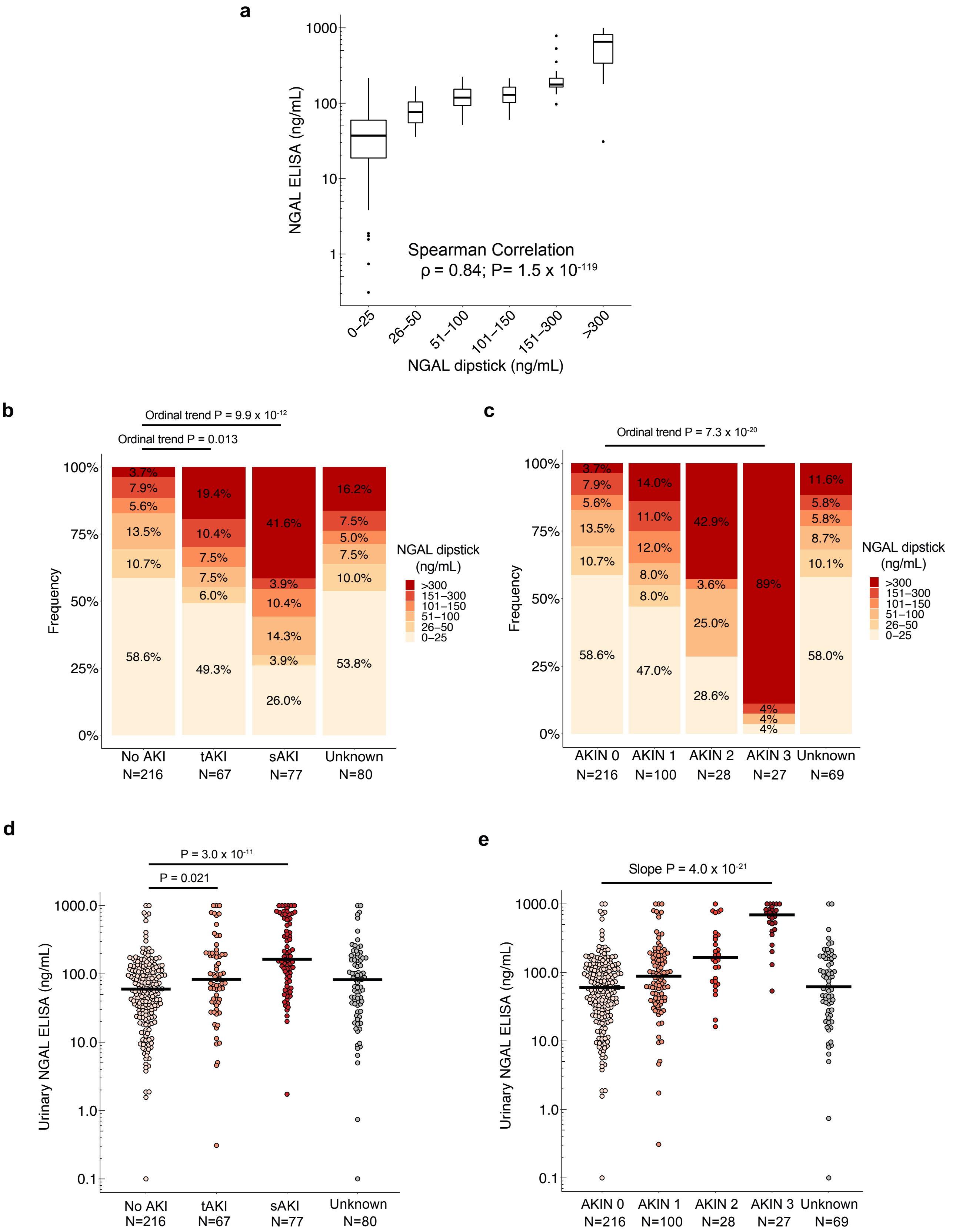


**Figure S2. Comparison of urine NGAL measurements by ELISA or Dipstick: (a)** Correlation between urine dipstick and ELISA measurements. **(b-c)** Urinary NGAL measured by dipstick is elevated by sustained AKI and dose-responsive to AKIN stage. **(d-e)** Urinary NGAL measured by ELISA is elevated by sustained AKI and dose-responsive to AKIN stage. Patients in the "Unknown" category (insufficient data for AKI or AKIN classification) are depicted, but excluded from statistical analyses. Diagnostic cutoffs for AKIN stage 2-3 are >150 ng/mL uNGAL (80% specificity, 75% sensitivity). Bars represent medians.

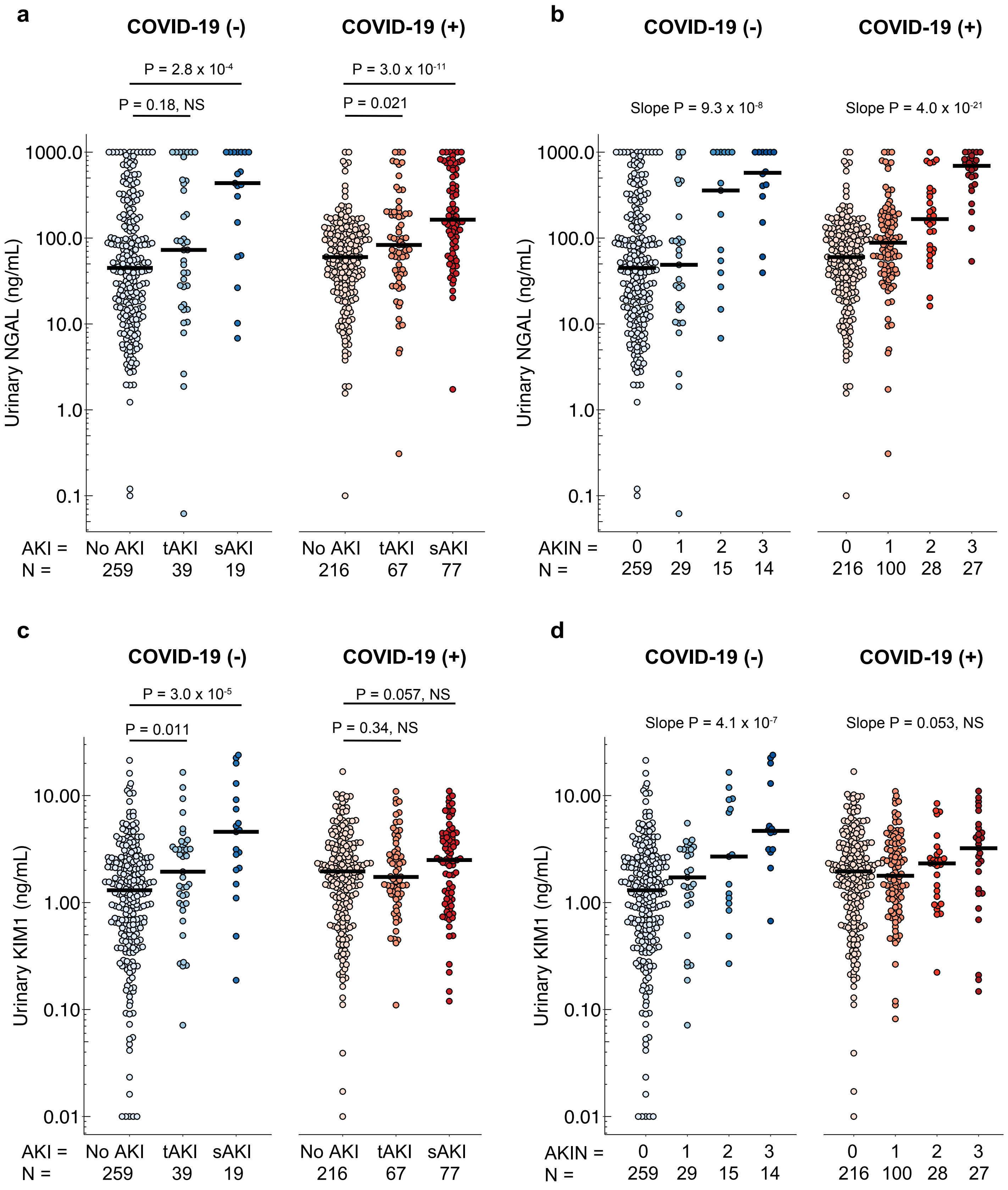


**Figure S3. Urinary NGAL and KIM-1 in COVID-negative and COVID-positive cohorts:** Urinary NGAL levels by **(a)** diagnosis and duration of AKI, and by **(b)** severity of AKI. **(c)** Urinary KIM-1 levels by diagnosis and duration of AKI, and by **(d)** severity of AKI. Bars represent medians. Note that in the setting of COVID-19, uKIM-1 was elevated in all patients, including those with AKIN stage 0 (no elevation of SCr).

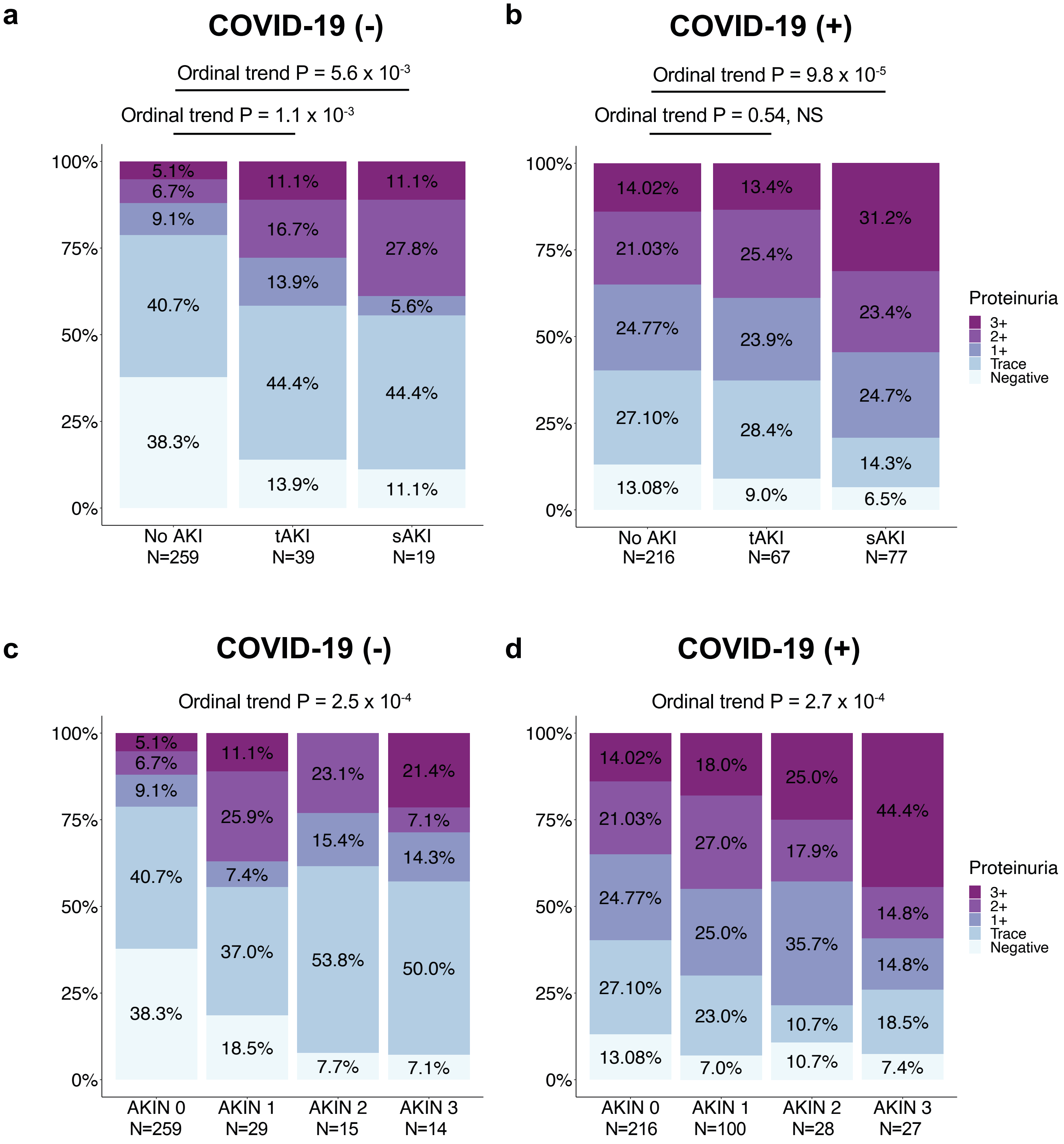


**Figure S4. Comparison of proteinuria in COVID-19-negative and COVID-19-positive cohorts by (a-b)** AKI diagnosis and duration; and by **(c-d)** increasing AKIN stage.

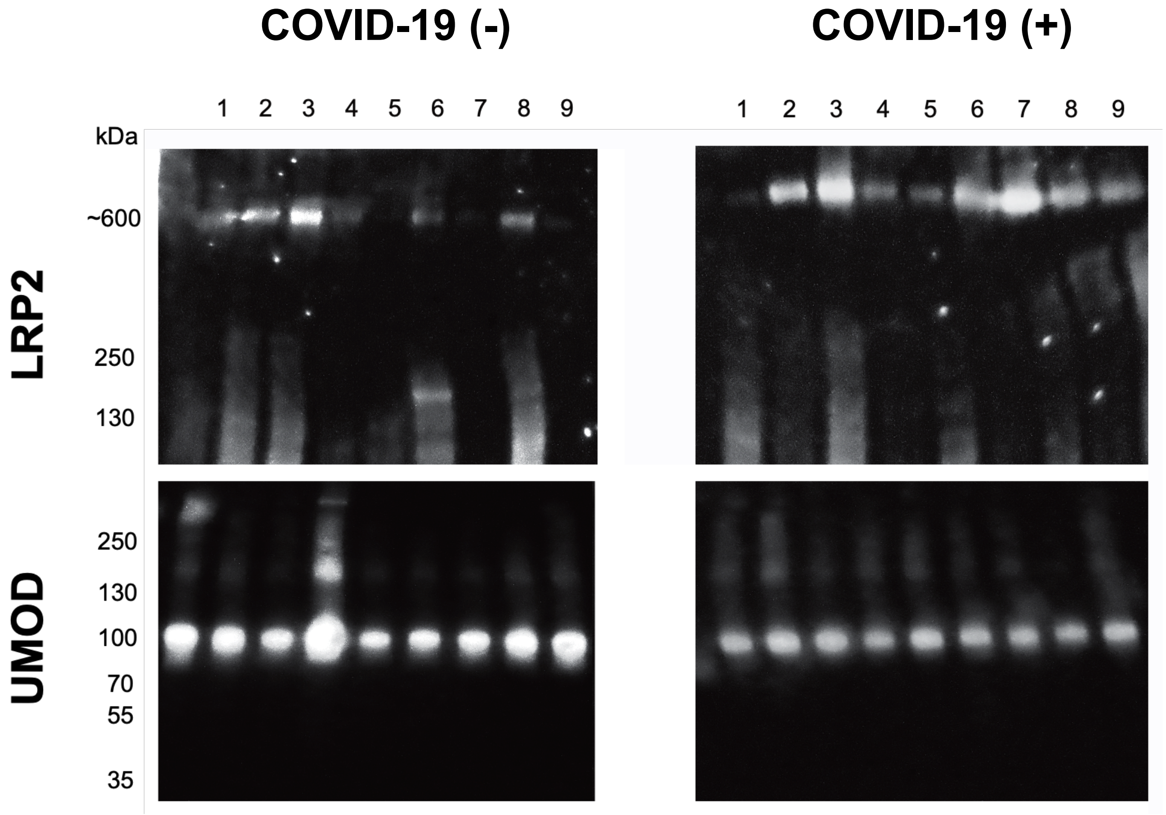


**Figure S5. Shedding of LRP2+ proximal tubule cells into the urine in COVID-19 cases:** Urine cell pellets from COVID-19 positive patients without AKI (AKIN=0) demonstrated prominent LRP2+ (megalin) protein compared with urine cell pellets from non-COVID-19 patients without AKI (AKIN=0) (**top panels**). In contrast, UMOD+ (uromodulin) cells were present regardless of COVID-19 status (**bottom panels**). Molecular weights of LRP2 and UMOD are 600 kDa and 100 kDa, respectively.

| Baseline Characteristics | COVID-19 (+) Cohort N=440 | COVID-19 (-) Cohort N=426 | P-value |
|---|---|---|---|
| Age (mean (SD)) | 63.7 (19.2) | 60.3 (17.9) | P < 0.05 |
| Sex (%) | | | |
| Male | 247 (56.1) | 244 (57.3) | NS |
| Female | 193 (43.9) | 182 (42.7) | |
| Race (%) | | | |
| White or Caucasian | 126 (28.6) | 145 (34.0) | NS |
| Black or African American | 90 (20.5) | 77 (18.0) | |
| Asian | 16 (3.6) | 11 (3.0) | |
| Unknown | 208 (47.2) | 193 (45.0) | |
| Ethnicity (%) | | | |
| Hispanic or Latinx | 238 (54.1) | 104 (24.0) | P < 0.05 |
| Non-Hispanic or Non-Latinx | 115 (26.1) | 94 (22.0) | |
| Unknown | 87 (19.8) | 228 (54.0) | |
| Presenting Comorbidities | | | |
| Obesity (%) | 188 (42.7) | NA | |
| Hypertension (%) | 143 (32.5) | NA | |
| Diabetes (%) | 88 (20.0) | NA | |
| Cardiovascular Disease (%) | 49 (11.1) | NA | |
| Pulmonary Disease (%) | 52 (11.8) | NA | |
| Cancer (%) | 62 (14.1) | NA | |
| Transplant (%) | 28 (6.4) | NA | |
| Chronic Kidney Disease (%) | | | |
| No CKD or CKD 1-2 | 234 (53.2) | 281 (66.0) | P < 0.05 |
| CKD 3-5 | 170 (38.6) | 145 (34.0) | |
| Kidney Transplant | 23 (5.2) | 0 (0.0) | |
| Unknown | 13 (3.0) | 0 (0.0) | |
| Baseline SCr (mean (SD)) | 1.24 (1.22) | 1.20 (0.91) | NS |
| Presenting Proteinuria (%) | | | |
| Negative | 57 (13.0) | 142 (33.4) | P < 0.05 |
| Trace | 107 (24.5) | 156 (36.7) | |
| 1+ | 107 (24.5) | 41 (9.6) | |
| 2+ | 94 (21.5) | 40 (9.4) | |
| 3+ | 72 (16.5) | 23 (5.4) | |
| Unknown | 0 (0.0) | 23 (5.4) | |
| Outcomes | | | |
| AKIN Stage (%) | | | |
| AKIN 0 | 216 (49.1) | 260 (61.0) | P < 0.05 |
| AKIN 1 | 100 (22.7) | 29 ( 6.8) | |
| AKIN 2 | 28 ( 6.4) | 15 (3.5) | |
| AKIN 3 | 27 ( 6.1) | 14 (3.3) | |
| Unknown | 69 (15.7) | 108 (25.4) | |
| AKI Status (%) | | | |
| No AKI (AKIN 0) | 216 (49.1) | 260 (61) | P < 0.05 |
| Transient AKI | 67 (15.2) | 39 (9.1) | |
| Sustained AKI | 77 (17.5) | 19 (4.5) | |
| Unknown | 80 (18.2) | 108 (25.4) | |
| Death within 90 days (%) | 117 (26.6) | 42 (9.9) | P < 0.05 |
| Dialysis (%) | 24 (5.5) | 9 (2.1) | P < 0.05 |
| Shock (%) | 162 (36.8) | NA | |
| Respiratory Failure (%) | 340 (77.3) | NA | |
| Length of Hospital Stay (mean (SD)) | 20.99 (22.96) | 6.86 (8.97) | P < 0.05 |

**Table S1. Baseline characteristics and outcomes for COVID-19 positive and negative cohorts:** The COVID-19 cohort includes all patients including “Unknown”. The COVID-19-negative cohort is a historical comparison cohort that we previously published in Stevens et al. Kidney Int Rep 2020. NS = not significant; NA = data not available.

| | Minimally-Adjusted | | | Fully-Adjusted-1 | | | Fully-Adjusted-2 (with Proteinuria) | | |
|---|---|---|---|---|---|---|---|---|---|
| **Urinary NGAL** | | | | | | | | | |
| **Primary Outcomes** | **OR** | **95% CI** | **P-value** | **OR** | **95% CI** | **P-value** | **OR** | **95% CI** | **P-value** |
| AKIN 1-3 | 2.36 | (1.78−3.21) | 1.04E-08 | 1.92 | (1.42−2.65) | 1.62E-04 | 1.89 | (1.39−2.63) | 8.80E-05 |
| AKIN 2-3 | 6.86 | (4.24−11.7) | 9.26E-14 | 6.89 | (3.99−12.63) | 4.16E-11 | 6.90 | (3.98−12.66) | 4.89E-11 |
| AKIN 3 | 24 | (10−71) | 3.09E-10 | 34 | (11−153) | 9.61E-08 | 37 | (11−169) | 1.12E-07 |
| Sustained AKI | 3.23 | (2.25−4.77) | 7.85E-10 | 2.67 | (1.81−4.06) | 1.80E-06 | 2.55 | (1.71−3.89) | 8.51E-06 |
| **Secondary Outcomes** | **OR** | **95% CI** | **P-value** | **OR** | **95% CI** | **P-value** | **OR** | **95% CI** | **P-value** |
| Death | 1.73 | (1.29−2.33) | 2.83E-04 | 1.62 | (1.19−2.24) | 2.53E-03 | 1.51 | (1.10−2.11) | 1.23E-02 |
| Dialysis | 4.92 | (2.75−9.34) | 2.68E-07 | 3.67 | (1.89−7.57) | 2.24E-04 | 3.59 | (1.83−7.45) | 3.33E-04 |
| Dialysis or Death | 2.38 | (1.77−3.26) | 2.52E-08 | 1.98 | (1.44−2.78) | 4.36E-05 | 1.84 | (1.33−2.61) | 3.71E-04 |
| Shock | 1.62 | (1.28−2.08) | 8.90E-05 | 1.64 | (1.26−2.15) | 2.91E-04 | 1.60 | (1.22−2.11) | 7.85E-04 |
| Shock or Death | 1.84 | (1.45−2.38) | 1.12E-06 | 1.79 | (1.38−2.35) | 2.28E-05 | 1.71 | (1.31−2.26) | 1.35E-04 |
| Respiratory Failure | 1.25 | (0.97−1.61) | 8.16E-02 | 1.39 | (1.05−1.86) | 2.31E-02 | 1.22 | (0.91−1.64) | 1.91E-01 |
| Respiratory Failure or Death | 1.50 | (1.15−1.96) | 2.89E-03 | 1.63 | (1.21−2.21) | 1.55E-03 | 1.43 | (1.05−1.95) | 2.40E-02 |
| **Time-to-Event** | **HR** | **95% CI** | **P-value** | **HR** | **95% CI** | **P-value** | **HR** | **95% CI** | **P-value** |
| 90-Day Mortality | 1.48 | (1.19−1.85) | 4.03E-04 | 1.40 | (1.11−1.77) | 5.09E-03 | 1.30 | (1.02−1.66) | 3.71E-02 |
| Length of Hospital Stay | 1.25 | (1.12−1.38) | 4.50E-05 | 1.22 | (1.09−1.36) | 4.80E-04 | 1.18 | (1.06−1.32) | 3.20E-03 |
| **Urinary KIM-1** | | | | | | | | | |
| **Primary Outcomes** | **OR** | **95% CI** | **P-value** | **OR** | **95% CI** | **P-value** | **OR** | **95% CI** | **P-value** |
| AKIN 1-3 | 1.16 | (0.92−1.48) | 2.08E-01 | 1.17 | (0.91−1.53) | 2.31E-01 | 1.13 | (0.87−1.48) | 3.77E-01 |
| AKIN 2-3 | 1.46 | (1.04−2.11) | 3.69E-02 | 1.35 | (0.94−1.99) | 1.20E-01 | 1.30 | (0.90−1.94) | 1.81E-01 |
| AKIN 3 | 1.62 | (1.01−2.72) | 5.93E-02 | 1.38 | (0.84−2.38) | 2.30E-01 | 1.32 | (0.79−2.32) | 3.05E-01 |
| Sustained AKI | 1.33 | (0.98−1.82) | 7.24E-02 | 1.32 | (0.95−1.86) | 1.09E-01 | 1.22 | (0.87−1.73) | 2.61E-01 |
| **Secondary Outcomes** | **OR** | **95% CI** | **P-value** | **OR** | **95% CI** | **P-value** | **OR** | **95% CI** | **P-value** |
| Death | 1.11 | (0.86−1.45) | 4.21E-01 | 1.09 | (0.84−1.44) | 5.25E-01 | 1.00 | (0.76−1.33) | 9.89E-01 |
| Dialysis | 1.49 | (0.93−2.53) | 1.22E-01 | 1.07 | (0.64−1.89) | 8.10E-01 | 1.00 | (0.59−1.79) | 9.85E-01 |
| Dialysis or Death | 1.17 | (0.92−1.51) | 2.09E-01 | 1.10 | (0.85−1.43) | 4.86E-01 | 0.99 | (0.76−1.31) | 9.54E-01 |
| Shock | 1.30 | (1.05−1.61) | 1.76E-02 | 1.27 | (1.02−1.60) | 3.70E-02 | 1.24 | (0.98−1.57) | 7.67E-02 |
| Shock or Death | 1.28 | (1.05−1.58) | 1.89E-02 | 1.27 | (1.03−1.59) | 3.06E-02 | 1.21 | (0.97−1.52) | 1.02E-01 |
| Respiratory Failure | 1.49 | (1.19−1.89) | 7.66E-04 | 1.47 | (1.15−1.90) | 2.28E-03 | 1.30 | (1.01−1.69) | 4.47E-02 |
| Respiratory Failure or Death | 1.48 | (1.17−1.89) | 1.33E-03 | 1.48 | (1.15−1.92) | 3.00E-03 | 1.30 | (0.99−1.70) | 5.61E-02 |
| **Time-to-Event** | **HR** | **95% CI** | **P-value** | **HR** | **95% CI** | **P-value** | **HR** | **95% CI** | **P-value** |
| 90-Day Mortality | 1.07 | (0.87−1.31) | 5.20E-01 | 1.06 | (0.87−1.30) | 5.57E-01 | 0.96 | (0.78−1.19) | 7.25E-01 |
| Length of Hospital Stay | 1.10 | (0.99−1.23) | 7.00E-02 | 1.11 | (0.99−1.23) | 7.00E-02 | 1.07 | (0.96−1.19) | 2.40E-01 |
| **Proteinuria** | | | | | | | | | |
| **Primary Outcomes** | **OR** | **95% CI** | **P-value** | **OR** | **95% CI** | **P-value** | **OR** | **95% CI** | **P-value** |
| AKIN 1-3 | 1.26 | (1.06−1.50) | 8.21E-03 | 1.16 | (0.96−1.39) | 1.29E-01 | NA | NA | NA |
| AKIN 2-3 | 1.33 | (1.05−1.70) | 1.87E-02 | 1.17 | (0.91−1.51) | 2.32E-01 | NA | NA | NA |
| AKIN 3 | 1.46 | (1.06−2.07) | 2.60E-02 | 1 | (1.00−2.00) | 3.58E-01 | NA | NA | NA |
| Sustained AKI | 1.45 | (1.18−1.81) | 6.42E-04 | 1.33 | (1.06−1.69) | 1.61E-02 | NA | NA | NA |
| **Secondary Outcomes** | **OR** | **95% CI** | **P-value** | **OR** | **95% CI** | **P-value** | **OR** | **95% CI** | **P-value** |
| Death | 1.33 | (1.10−1.61) | 3.48E-03 | 1.30 | (1.07−1.59) | 9.74E-03 | NA | NA | NA |
| Dialysis | 1.55 | (1.10−2.23) | 1.47E-02 | 1.26 | (0.86−1.86) | 2.40E-01 | NA | NA | NA |
| Dialysis or Death | 1.45 | (1.21−1.74) | 7.60E-05 | 1.34 | (1.10−1.63) | 3.38E-03 | NA | NA | NA |
| Shock | 1.17 | (1.00−1.37) | 5.55E-02 | 1.15 | (0.97−1.36) | 1.04E-01 | NA | NA | NA |
| Shock or Death | 1.26 | (1.08−1.47) | 3.67E-03 | 1.22 | (1.04−1.44) | 1.56E-02 | NA | NA | NA |
| Respiratory Failure | 1.58 | (1.30−1.95) | 8.22E-06 | 1.65 | (1.33−2.07) | 8.98E-06 | NA | NA | NA |
| Respiratory Failure or Death | 1.69 | (1.37−2.12) | 1.88E-06 | 1.71 | (1.36−2.17) | 6.98E-06 | NA | NA | NA |
| **Time-to-Event** | **HR** | **95% CI** | **P-value** | **HR** | **95% CI** | **P-value** | **HR** | **95% CI** | **P-value** |
| 90-Day Mortality | 1.29 | (1.11−1.49) | 6.16E-04 | 1.26 | (1.09−1.47) | 2.36E-03 | NA | NA | NA |
| Length of Hospital Stay | 1.18 | (1.09−1.29) | 1.30E-04 | 1.17 | (1.07−1.29) | 5.00E-04 | NA | NA | NA |

**Table S2. Urinary biomarker associations with primary and secondary outcomes:** minimally-adjusted model includes age, sex, race, ethnicity as covariates; fully-adjusted-1 model includes age, sex, race, ethnicity, baseline SCr, pre-existing obesity, diabetes, hypertension, solid or hematologic transplant, cancer, cardiovascular disease, and pulmonary disease; fully-adjusted-2 (with proteinuria) model includes proteinuria from the same urine sample along with all of the covariates from the fully-adjusted model-1; log-transformed and standard normalized urinary biomarkers were tested as continuous predictors of binary outcomes using logistic regression. Cox-proportional hazards regression model was used for 90-day mortality analysis. Competing risks regression was used to derive hazards ratios for length of hospital stay with death as a competing risk. Odds ratios (OR) and hazard ratios (HR) are expressed per one unit of biomarker standard deviation.
